# Social and private value created through commercialization of Gilead Sciences’ innovative medicines for Hepatitis C and HIV: a cross-sectional analysis

**DOI:** 10.1101/2025.06.11.25329435

**Authors:** Paula G. Chaves da Silva, Rena M. Conti, Fred D. Ledley

## Abstract

**Objectives:** This work assesses the value created for society and for private sector by Gilead Sciences, Inc. (Gilead) through commercialization of medicines for hepatitis C and HIV from 2012-2020. While Gilead has developed several groundbreaking medicines, the company has also been criticized for its drug pricing, patenting, and profits. This analysis posits the total value created by a medicine derives from the health benefit provided to those using these medicines.

**Design:** Cross-sectional analysis.

**Primary and Secondary outcome measures:** Health benefit from treatment for nine hepatitis C or HIV medicines (Quality-Adjusted Life Years [QALYs], $), number of individuals benefited, price paid, and Gilead Sciences’ financial report (10K).

**Results:** Gilead’s product sales generated 15.1 million in health benefit measured using QALYs for 23.7 million people. Expressed in monetary terms, these products created a total health value of $744.8 billion (US: $168.0; global: $576.8). This produced a residual health value, net price paid, of $544.6 billion (US: $35.9; global: $508.7) including $418.6 billion from hepatitis C medicines (US: $62.4; global: $356.2) and $125.9 billion from HIV medicines (US: -$26.6; global: $152.5). Total social value (US + global) created was $613.6 billion including residual health value ($544.6 billion), R&D ($34.2 billion), job creation ($9.7 billion), and social payments ($25.1 billion). Total private value was $193.5 billion including shareholder value ($85.7 billion) and network value (payments to commercial firms, $107.8 billion).

**Conclusions:** Social and private value differed between US and global sales and among diseases. This analysis highlights the scale of the value that can be created by modern medicines and the commercialization of pharmaceutical products applied to unmet medical needs. Assessing value creation at product level may guide corporate strategies and inform policies to promote both profit and public purpose.

**Strengths and limitations of this study:**

- “Total stakeholder value” approach considers the sum of value provided to different stakeholders
- Total health value measured from unit sales and health benefit to treated individuals
- Analysis based on market data and financial reports of manufacturers applicable to business strategy and policy
- Method applicable to assessing value created by individual drugs, classes of drugs, or companies
- Health benefit measured in QALYs are subject to limitations of this metric

**MeSh Headings:** “Hepatitis C” [MeSh Terms], “HIV Infections” [MeSh Terms], “Quality-Adjusted Life Years” [MeSh Terms], “Private Sector” [MeSh Terms], “Public Sector” [MeSh Terms]

## INTRODUCTION

Commercialization of new medicines is expected to create value for patients, society, and pharmaceutical firms. This study assesses the value created through commercialization of a series of innovative medicines developed by Gilead Sciences, Inc. (Gilead) including groundbreaking cures for hepatitis C (HCV), first-line treatments for HIV, and the first products for HIV pre-exposure prophylaxis (PrEP). While several of these medicines are recognized as “essential medicines” by the World Health Organization (1), Gilead has faced intense criticism and congressional scrutiny for its business practices regarding drug pricing, patenting, executive compensation, and distributions to shareholders (2-7), and has been portrayed as prioritizing private interests over those of society (5, 6, 8).

This work assesses the total value created through commercialization of Gilead’s HCV and HIV products as well as the balance between social and private value creation. The analysis utilizes a “total stakeholder value” approach (9, 10) which considers the sum of value provided to different stakeholders. The premise of this analysis is that the totality of the value created by a medicine derives from the health benefit provided to those utilizing the product (total health value). This value is then distributed among various stakeholders through the price paid for the drug and industry’s use of the proceeds from product sales. This generates different forms of value (11-13) including residual health value (net price paid) for those treated with the medicine, profits for shareholders (shareholder value), jobs for employees (job creation), funding for scientific and technological advances (scientific value), an expanded tax base, support for public institutions (social payments), and revenue for a network of suppliers, contractors, or distributors in the biopharmaceutical supply chain (network value) (14). In this analysis, these different forms of value are categorized as either “social” or “private” value as shown in figure 1.

**Figure 1.**
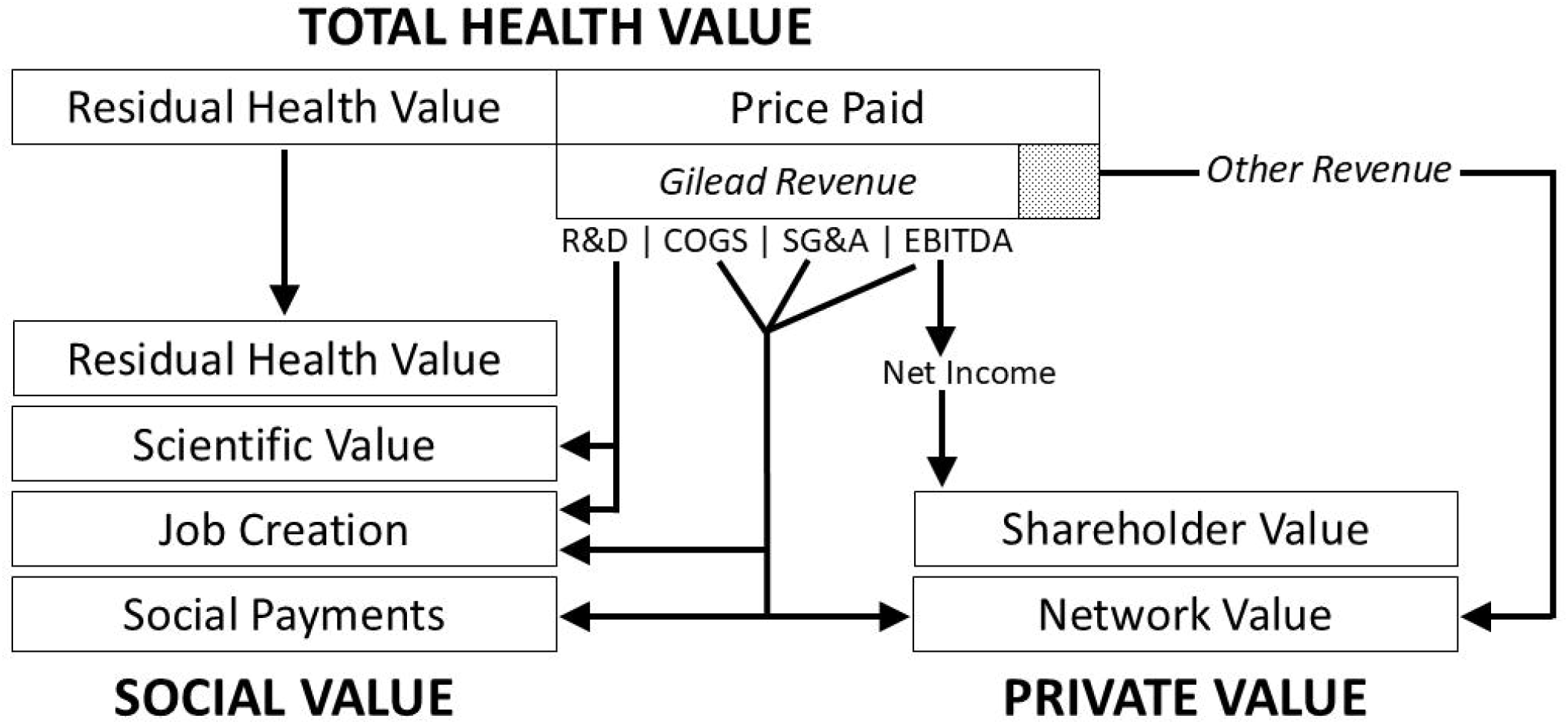
Value creation from the commercialization of Gilead Sciences’ medicines for HCV and HIV. This analysis posits that the value created by a new medicine is embodied in the total health value (health benefit) provided to those using the medicine. This value is then distributed between social value and private value by the price paid for the product and how these revenues are expensed for different purposes. Gilead revenue is reported net of rebates. In this framework, the social value created includes the residual health value (net price paid) along with the scientific value generated by R&D, the value of jobs created, and social payments to public sector organizations. The private value created includes shareholder value (change in market capitalization plus cash distributions to shareholders) as well as payments made to other private sector organizations.

Social value has been defined as the *“…benefits or reductions of costs for society… that go beyond the private gains and general benefits of market activity*” (15) or, in the context of corporations, the value accruing to “…*non-investor stakeholders affected by business: individuals, employees, communities, and society*” (10). In this study, residual health value, scientific value, job creation, and payments to government or other public sector organizations (social payments) are considered social value.

Private value typically comprises the value accruing to the producer’s shareholders (“shareholder value”), which derives from changes in the company’s stock price as well as the distribution of corporate profits to shareholders through dividends or stock buybacks (5). In this study, private value also includes the amounts paid by the manufacturer to other firms that provide goods, services, or contract operations (“network value”) (14, 16).

The health benefit created through commercialization of a medicine is estimated as the number of individuals prescribed that medicine times the average benefit received measured in quality-adjusted life years (QALYs) (17-20). The value of this health benefit in monetary terms is estimated as the health benefit in QALYs times a globally adjusted measure of customer’s willingness to pay (WTP) for a year of healthy life (i.e., one QALY) or WTP/QALY (21). This analysis considers the residual health value (net price paid) to be the most appropriate measure of value accruing to those treated with a medicine. This reflects evidence that high drug prices are associated with poor compliance of prescribed treatment regiments as well as greater economic, food, and housing insecurity, which may collectively compromise the ability of individuals to realize the full benefit of Gilead’s HCV and HIV medicines (22-24).

Unlike traditional economic methods, this approach assesses the value created through commercialization of individual medicines, classes of medicines, or companies based on sales and pricing data for individual drugs and audited corporate financial reports. As such, these findings could guide strategic management of innovation in the biopharmaceutical industry, aiming to address the needs of both shareholders and society (10, 25, 26) while supporting policies designed to promote innovation for unmet burdens of disease and achieve both profit and public purpose.

### Patient and public involvement

Patients and the public were not involved in this study.

## MATERIALS AND METHODS

### Data sources

Data sources and exclusion criteria are reported in detail in supplemental methods 1. Briefly, Gilead’s marketed products for HCV or HIV were identified in annual reports. Units sold/year and the wholesale price paid for US and 77 global markets was from IQVIA MIDAS®. IQVIA MIDAS™ integrates IQVIA’s national audits into a globally consistent view of the pharmaceutical market, tracking products in hundreds of therapeutic classes, and provides estimated product volumes, trends and market share through retail and non-retail channels. MIDAS data is updated monthly and retains 12 years of history.

QALY data for Gilead products were from the Tufts Center for Evaluation of Value and Risk (CEVR) database or extracted through manual literature review. Gilead’s annual financial data were obtained from Compustat (WRDS).

### Estimating health value

The analysis is illustrated in figure 1. The health benefit created by medicine “m” in year “y” was calculated in QALYs as:

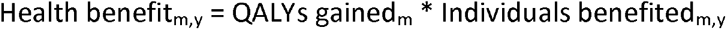

where QALYs are estimated based on the health outcome relative to no treatment. The number of individuals benefited was estimated as the total price paid divided by units sold/year considering a 90-pill treatment for HCV and an average 260 pills/treatment for HIV. For PrEP, individuals benefited is the number individuals benefited divided by “Number Needed to Treat” (NNT)= 58.1 (27).

QALYs gained was the median of QALYs treated minus QALYs untreated from publications reporting both values (supplemental table 1). While this work uses QALY units as a measure of health value, there are essential differences in our use of this metric and its application in cost-effectiveness studies as discussed in detail in supplemental methods 1. QALYs are typically reported as the lifetime health benefit provided by a medicine’s indicated use. For HCV, one year of therapy provides lifetime benefit. For HIV, one year of therapy provides one year of (annual) benefit converted from lifetime QALYs using the geometric series:

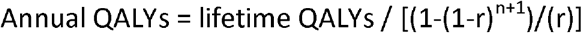

where r = discount rate for future health benefits, and n = number of years (time horizon) for the reported data.

Health value in USD was calculated from health benefit using a globally adjusted (“overall predicted mean”) value for WTP/QALY ($52,619.40) that accounts for differences in methodology and respondent characteristics through econometric meta-regression (21). Health value was also estimated using an averaged US WTP/QALY of $104,000 (28) (supplemental table 2).

Total health value was the sum of the health value created by each of the nine medicines for HCV or HIV 1/2012-6/2020. Dollars are inflation-adjusted to 2016.

### Estimation of social and private value creation

Social value in year y was calculated as

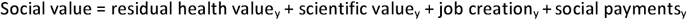

where residual health value = total health value minus price paid; scientific value = R&D spending; job creation = number of employees * average industry salary (29); and social payments include taxes, fines, donations and reported payments to public institutions.

Private value in year y was calculated as:

Private value = shareholder value_y_ + network value_y_

Shareholder value in year y was calculated as:

Shareholder value = change in market capitalization_y_ + dividends_y_ + stock buybacks_y_ where change in market capitalization is the market capitalization in year y minus the market capitalization in the previous year. Network value in year y was calculated as:

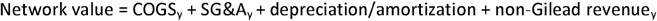

where non-Gilead revenue = the price paid for the product minus Gilead’s product-specific revenue. This includes revenue to firms selling unbranded products, intermediates in the distribution network including suppliers, contractors PBMs, retailers, distributors, or others, and payments not recognized by Gilead as revenue (16).

## RESULTS

### Gilead products for HCV and HIV

Table 1 shows published data on QALYs gained (health benefit) from the use of nine Gilead medicines meeting the inclusion criteria (supplemental methods 1, supplemental tables 1, 3, 4). These products account for 77.6% of Gilead revenue 2012-2020 (supplemental table 4). QALYs are estimated relative to those receiving no treatment or placebo and were expressed either as a lifetime value or annual value. For HCV medicines, median lifetime QALYs gained varied from 2.60 QALYs for Sovaldi to 3.94 for Epclusa. For HIV medicines, Truvada (non-PrEP), Descovy (non-PrEP), Atripla, Stribild, Genvoya, and Biktarvy, median annual QALYs gained was 0.23 QALYs. For PrEP, Descovy or Truvada, median annual QALYs gained was 0.35 (supplemental table 1).

**Table 1.**
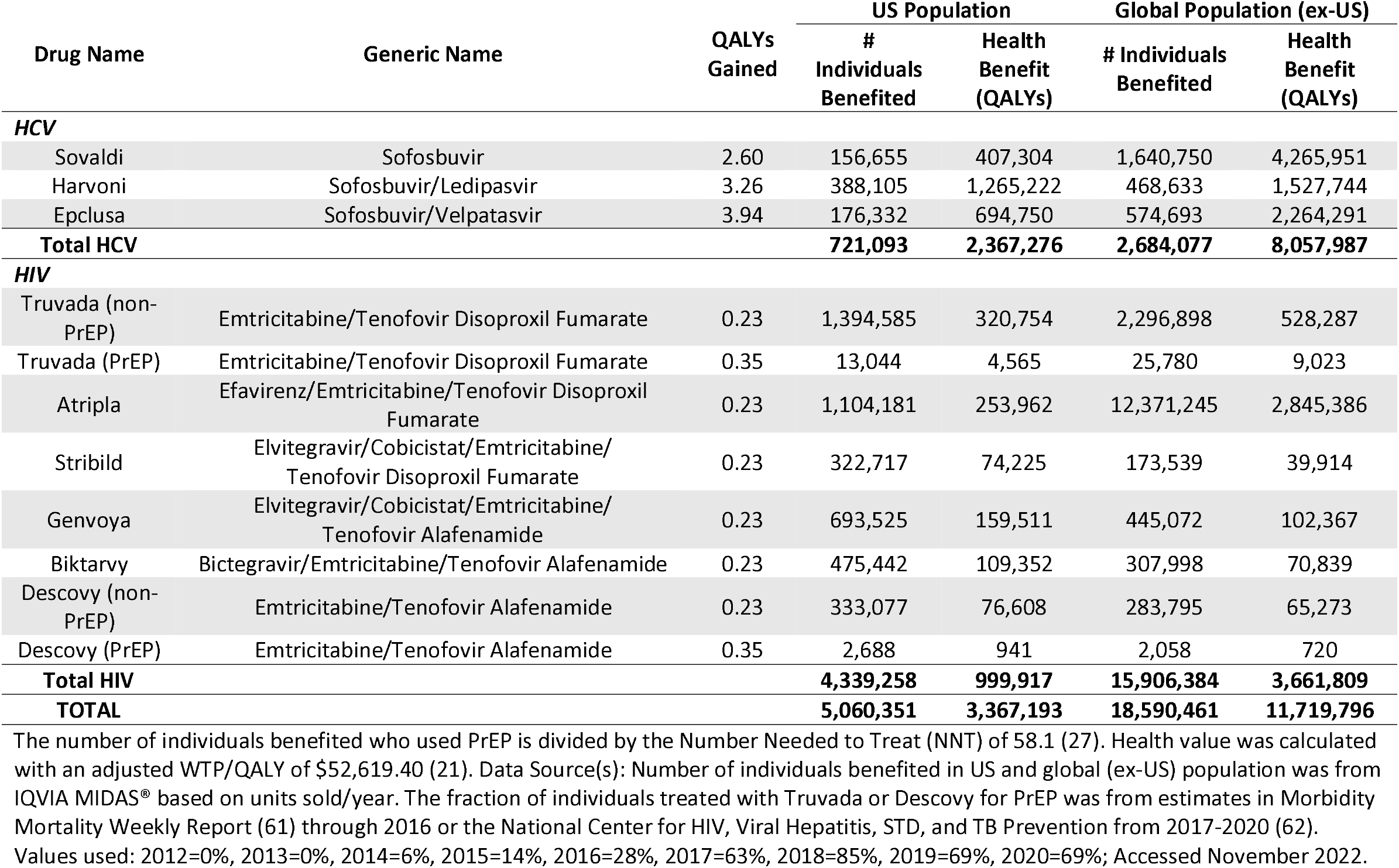
Total health benefit created by Gilead’s medicines 2012-2020 in QALYs.

### Total health value created by Gilead HCV and HIV medicines 2012-2020

Table 1 shows the total health benefit, created by these medicines expressed in QALYs. Table 2 shows the total health value calculated from the health benefit in USD using an adjusted average WTP/QALY of $52,619.40 (21). The total health value created by US sales of HCV medicines was $118.0 billion, with Harvoni generating the greatest sales and health value. Global (ex-US) sales of HCV medicines generated total health value of $396.1 billion led by Sovaldi with Sovaldi also generating the greatest health value.

**Table 2.** Total and residual health value created by Gilead’s medicines 2012-2020 in USD US Population Global Population (ex-US)

| Generic Name | US Population |  |  | Global Population (ex-US) |  |  |
| --- | --- | --- | --- | --- | --- | --- |
|  | Price Paid | Health Value | Residual Health Value | Price Paid | Health Value | Residual Health Value |
| **HCV** |  |  |  |  |  |  |
| Sovaldi | \$13.1 | \$20.8 | \$7.7 | \$13.0 | \$211.2 | \$198.2 |
| Harvoni | \$33.4 | \$63.7 | \$30.2 | \$15.9 | \$75.9 | \$60.0 |
| Epclusa | \$9.0 | \$33.6 | \$24.6 | \$10.9 | \$108.9 | \$98.0 |
| **Total HCV** | **\$55.5** | **\$118.0** | **\$62.4** | **\$39.8** | **\$396.1** | **\$356.2** |
| **HIV** |  |  |  |  |  |  |
| Truvada | \$22.2 | \$16.6 | -\$5.6 | \$11.3 | \$27.3 | \$16.0 |
| Atripla | \$17.3 | \$13.0 | -\$4.3 | \$8.8 | \$139.9 | \$131.0 |
| Stribild | \$6.7 | \$3.7 | -\$2.9 | \$1.5 | \$2.0 | \$0.54 |
| Genvoya | \$14.8 | \$7.7 | -\$7.0 | \$3.5 | \$4.9 | \$1.5 |
| Biktarvy | \$10.5 | \$5.2 | -\$5.4 | \$1.4 | \$3.3 | \$1.9 |
| Descovy | \$5.2 | \$3.8 | -\$1.4 | \$1.8 | \$3.2 | \$1.5 |
| **Total HIV** | **\$76.6** | **\$50.0** | **-\$26.6** | **\$28.2** | **\$180.7** | **\$152.5** |
| **TOTAL** | **\$132.2** | **\$168.0** | **\$35.9** | **\$68.1** | **\$576.8** | **\$508.7** |

All values are in billions and are inflation-adjusted to 2016. Price paid: wholesale price paid net of prompt pay discounts and chargebacks. Health Value: The overall health value attributed to all products combined. Health value was calculated with an adjusted WTP/QALY of \$52,619.40 (21). Residual Health Value: For the US, the net health value was calculated as the difference between the total US health value generated by the product and its spending within the US. The same calculation applies to Global (excluding the United States). A positive value indicates a net health gain, while a negative value suggests a net health loss. Data Source(s): IQVIA MIDAS®, Centers for Medicare & Medicaid Services (CMS), and AtlasPlus Centers for Disease Control and Prevention (CDC); Accessed May 2022.

The total health value created by US sales of HIV medicines was $50.0 billion with the largest fraction generated by Truvada (non-PrEP) sales. The total health value created by global sales of HIV medicines was $180.7 billion with the largest fraction generated by Atripla. Truvada or Descovy sales (PrEP) generated proportionally less health value.

Together, sales of HCV and HIV medicines created total US health value of $168.0 billion and total global health value of $576.8 billion with greater global health value generated by HCV medicines despite lower sales than US HCV medicines.

The US and global total health value was also calculated with the US WTP/QALY in supplemental table 2.

### Residual health value of HCV and HIV medicines 2012-2020

Table 2 shows the residual health value generated by these medicines. Sales of these medicines 2012-2020 totaled $132.2 billion in the US and $68.1 billion globally including sales of Gilead’s branded, authorized generic products, and unbranded products licensed by Gilead (30). In the US, the residual health value of these medicines was $35.9 billion (21.3% of total US health value) including $62.4 billion from HCV medicines and -$26.6 billion for HIV medicines. Globally, the residual health value was $508.7 billion (88.2% of total global health value. Both HCV and HIV medicines had positive residual health values – $356.2 billion for HCV and $152.5 billion for HIV.

The US and global residual health value was also calculated with the US WTP/QALY in supplemental table 5.

### Distribution of Gilead product revenues 2012-2020 to social and private value

Gilead’s revenues from these products, net of rebates, refunds, credits, and returns (supplemental figure 1), peaked at $33.1 billion in 2015 and totaled $201.0 billion 2012-2020 (supplemental table 6). Net income (earnings) paralleled revenues, increasing from $2.7 billion in 2012 to $18.3 billion in 2015 before dropping more than tenfold to $114.1 million in 2020 (supplemental table 6, supplemental figure 2-A). Gilead’s market capitalization paralleled their revenue and net income, increasing from $58.3 billion in 2012 to $145.7 billion in 2015 before dropping to $67.7 billion in 2020 (supplemental figure 2-B). From 2012-2020, distributions to shareholders through dividends or stock buybacks totaled $50.8 billion (78% of earnings). Annual change shareholder value was $26.1 billion at the end of 2012, peaked at $61 billion in 2013 and was -$38.4 billion in 2016 (supplemental table 6), resulting in a total of $85.7 billion from 2012-2020.

Gilead’s cost of goods sold varied from $2.4 to $3.3 billion. SG&A increased from $1.2 billion in 2012 to $4.2 billion in 2020 (supplemental table 6, supplemental figure 2-C). Overall, commercialization of these medicines generated $107.8 billion in network value 2012-2020 (table 3) including non-Gilead revenue of $44.2 billion.

**Table 3.** Components of social and private value generated by Gilead 2012-2020.

| Components of Value | Social Value | Private Value |
| --- | --- | --- |
| US Residual Health Value | \$35.9 | - |
| Global Residual Health Value | \$508.7 | - |
| Scientific Value | \$34.2 | - |
| Job Creation | \$9.7 | - |
| Social Payments | \$25.1 | - |
| Shareholder Value | - | \$85.7 |
| Network Value | - | \$107.8 |
| **TOTAL** | **\$613.6** | **\$193.5** |

All values are in USD (billions) and inflation-adjusted to 2016. Data Source(s): Authors’ analysis of data in Tables 1 and 2.

Gilead’s reported expenses contributed $25.1 billion to social payments 2012-2020. R&D expenses rose from $1.8 billion in 2012 to $4.7 billion in 2020 with a discontinuous peak of $7.8 billion in 2019 related to a research agreement with Galapagos NV. R&D expense totaled $34.2 billion 2012-2020. Gilead’s tax payments rose from $1.1 billion in 2012 to $3.6 billion in 2016 with a discontinuous peak of $8.7 billion in 2017 reflecting repatriated earnings of foreign subsidiaries. Taxes were negative in 2019 and totaled $24.5 billion 2012-2020. Gilead’s employment was 81,500 person years from 2012-2020 with an estimated total value of $9.7 billion. Other contributions to social value included $97.2 million in fines and $453.5 million paid to public sector institutions excluding in-kind contributions of product, service, or expertise, charitable contributions, and contributions (table 3).

### Comparing social and private value created

The social value created 2012-2020 was $613.6 billion including residual health value (US = $35.9 billion + global = $508.7 billion), scientific value (R&D = $34.2 billion), job creation ($9.7 billion), and social payments ($25.1 billion) (table 3, supplemental figure 3). The total private value created was $193.5 billion, estimated as the sum of shareholder value ($85.7 billion) and network value ($107.8 billion). The discrepancy between total health value ($744.8 billion) and the sum of social and private value ($807.1 billion) reflects both that the drugs considered in this scenario only accounts for 77.6% sales and that shareholder value is determined by market forces and not directly from revenue or profit.

## DISCUSSION

These data suggest that Gilead’s commercialization of medicines for HCV and HIV generated more than 15 million QALYs of health benefit for more than 23 million people globally from 2012-2020. Expressed in monetary terms, these products created a total health value of approximately $750 billion. The scale of the health benefit created by these medicines is a reminder of the large unmet, global burden of disease (in 2019 the global burden of HCV was 15.3 million DALYs (31) and HIV was 43.6 million DALYs (32)) as well as the healing potential of modern medicines and ecosystem for innovation and commercialization of pharmaceutical products.

This analysis also shows that the value created through commercialization of these products was disproportionately distributed to social value (76%) rather than private value (24%). Most of this social value was in the form of the residual health value provided to those treated with the products, with smaller contributions in the form of social payments, scientific value, or job creation. The majority of the private value created was in the form of network value (56%) with less in the form of shareholder value (44%). It should be noted that Gilead created little shareholder value after 2015. From 2015 to 2020, Gilead’s net income (earnings) dropped 99% and its market capitalization dropped 46% due to both increasing market competition for its products and a series of unproductive acquisitions or partnerships with Forty Seven, Nimbus Apollo, and Galapagos as well as significant write-offs from its acquisition of Kite Pharma. Gilead was able to preserve shareholder value over this period by distributing its profits to shareholders through dividends and stock buybacks.

The present results are broadly consistent with economic studies associating broad measures of pharmaceutical innovation with improvements in public health. Those studies demonstrate that the consumer surplus from pharmaceutical innovation (analogous to social value) is often substantially greater than the producer surplus (analogous to private value) (33-40). Our results are also broadly consistent with the findings of a recent study suggesting that 90% of the value generated by direct acting antiviral drugs for HCV resulted in health benefits to patients or cost-savings to society (consumer surplus) as opposed to revenues to pharmaceutical manufacturers (producer surplus) (40).

Our analysis differs from previous studies in that it focuses on the health value provided to individuals based on market data describing the number of individuals benefited and sales prices, considers the value accruing not only to pharmaceutical manufacturers, but other companies in the value network, and characterizes corporate revenues, spending, profit, and value creation using standard financial metrics and audited financial data. As such, the present observations are directly applicable to formulating corporate strategies that consider both responsibility to shareholders and to society (10, 25, 26) as well as public policies that balance the affordability of innovative medicines with incentives for innovation.

This analysis also shows that Gilead’s products generated substantially greater total health value and social value for global markets than for US markets. This was primarily due to the higher prevalence of HCV and HIV in global (ex-US) populations and larger number of individuals benefited. Global markets, however, also provided proportionally greater residual health value and proportionally greater social value due to the availability of less expensive, unbranded products outside of the US and Europe (30, 41). These low-price products increased social value creation both by enabling greater market penetration (42) and distributing a smaller fraction of total health value to the private sector.

The balance between social and private value creation was different for HCV and HIV medicines. While the HIV medicines accounted for >80% of individuals treated with Gilead products and the majority of Gilead’s revenues, HCV medicines contributed more than two thirds of the total health value and social value. This discrepancy reflects, in part, the greater health benefit associated with HCV medicines, which provides a lifetime cure from a short course of therapy, and a positive residual health value at 2012-2020 prices. In contrast, treatment for HIV extends and improves the quality of life only while the medicine is being administered, and pre-exposure prophylaxis (PrEP) requires treatment of 58.1 individuals to avert one new HIV infection. This resulted in a negative residual health value at 2012-2020 US prices and a net negative contribution to social value (43).

Finally, the value created through commercialization of these drugs represents a return on the public and private investments necessary to bring these medicines to the public (24, 44-49). The NIH spent at least $6.5 billion for research on chemistries for nucleic acid pharmaceuticals and the biology of the hepatitis C virus that informed discovery and development of HCV drugs (50, 51). The NIH also spent tens of billions of dollars for research on the HIV virus that informed discovery and development of HCV drugs (52). Many of the products considered in this analysis originated in companies founded by scientists from Emory University’s Center for Drug Discovery including emtricitabine, originated by Triangle Pharmaceuticals, which Gilead acquired for $464 million in 2022, and sofosbuvir, originated by Pharmasset, which Gilead acquired for $11 billion in 2011. In both cases, Gilead subsequently invested in phase 3 clinical trials, process development, and the regulatory submissions that led to marketing approval. While further research would be required to detail all the investment in these products, this analysis suggests that the scale of both social and private value created through commercialization of these products was sufficient to provide robust returns on both public and private investment.

These results do not obviate concerns regarding the availability or affordability of Gilead’s products. While the overall value created by Gilead’s product portfolio is weighed towards greater social value, this is not true for all Gilead products or for all patient populations. Specifically, Gilead’s products for HIV generated a net negative residual health value at prevailing US prices, despite generating a positive balance between social and private value creation globally. Moreover, the proportionally greater social value generated by HCV medicines was likely influenced by the intense public pressure on Gilead (7) and the work of patient advocacy groups, not-for-profit organizations, and public policies undertaken to improve access to these medicines through pricing, financing, licensing, capacity building, or donation (53-55).

## Limitations

There are several limitations to this study. First, use of QALY metrics for health technology assessment has been criticized (56) for inconsistent methodologies and biases related to disease severity, chronicity, and age (57). Our results, insofar as they utilize QALYs, may be subject to some of the same limitations. Second, estimates of WTP and WTP/QALY are known to be influenced by GDP, study design, and demographics (28, 58, 59). The globally adjusted average WTP/QALY derived by meta-regression (21) used in this analysis may not fully account for these factors.

Third, financial data reported using Generally Accepted Accounting Principles (US GAAP) reflects accrual-based accounting rather than actual cash flows. Product revenue is reported net of rebates, refunds, credits, and returns and is recognized when the product is delivered, not when revenues are received. Expenses do not include capitalized (investment) costs or donations of goods or services. Fourth, Gilead’s financial data, derived from SEC filings, describe only transactions considered material to the company’s stock price (60), may underrepresent small transactions, donations, or payments to public institutions leading to an underestimation of social value, and do not itemize employee costs.

Fifth, this analysis was limited by the lack of transparency regarding medicine utilization and sale prices. Utilization data from IQVIA/MIDAS does not include products donated through government or non-government organizations, sales outside traditional wholesale outlets or retail pharmacies, bundling or other price breaks off wholesale prices.

## CONCLUSION

Commercialization of Gilead Sciences’ HCV and HIV medicines 2012-2020 created more social than private value with the greatest social value reflecting health benefits provided by unbranded products for HCV in global markets. The balance between social and private value was impacted by the disease indication, corporate practices, and public policy. Assessing both the social and private value created by new medicines may inform public policies intended address the needs and expectations of all stakeholders in pharmaceutical innovation.

## ACKNOWLEDGMENTS

**Acknowledgements:** The authors thank Juliana Harrison, M.B.A., Program Manager, Bentley University for assistance preparing the manuscript; Xingqi Ye, Visiting Student, Cornell University for assistance collecting QALY data from the literature; Michael Boss, Ph.D., Executive in Residence, Bentley University, Nancy Hsiung, Ph.D., Executive in Residence, Bentley University, and Bruce Leicher, Esq., Executive in Residence, Bentley University, for their critical feedback.

## Article Information

### Contributors

FDL contributed as corresponding author. PGC and FDL designed the study. PGC, RMC and FDL analyzed and interpreted the data and performed the statistical analyses. PGC, RMC and FDL drafted the initial manuscript. PGC, RMC and FDL reviewed the manuscript and approved the final version of the manuscript. FDL is the guarantor and accepts full responsibility for the work and/or the conduct of the study, had access to the data, and controlled the decision to publish.

## Funding/Support

This work was supported by a grant from the National Biomedical Research Foundation to Bentley University (Grant number: N/A). The sponsors had no role in the design and conduct of the study; collection, management, analysis, and interpretation of the data; preparation, review, or approval of the manuscript; or the decision to submit the manuscript for publication. The sponsors did not have the right to veto publication or to control the decision regarding to which journal the paper was submitted.

## Competing Interests

The authors report no competing interests. Dr. Ledley reported receiving research funding from the National Biomedical Research Foundation, West Health Policy Center, Institute for New Economic Thinking, National Pharmaceutical Council, and Eastern Research Group since 2020. Dr. Conti served as a special economic consultant to the Center for Medicare and Medicaid Services in 2022-2023 and was a voting committee member of ICER in 2022-2024 unrelated to her efforts on this project. Dr. Conti is currently serving as a board member of BUMG and the State of New Jersey Drug Affordability Council. Dr. Conti served as a member of the state of Illinois Advisory Council on Financing and Access to Sickle Cell Disease Treatment and Other High-Cost Drugs and Treatment in 2024. During the period this project was ongoing, Dr. Conti’s research was funded by grants from the National Science Foundation, the Sloan Foundation, the Leukemia & Lymphoma Society, the National Cancer Institute, the National Institute for Drug Abuse, the Veteran’s Affairs Administration, the Department of Defense and Arnold Ventures. None of these granting agencies funded her efforts in this project. Dr. Conti serves as a consulting expert to Greylock McKinnon Associates and Keystone Consulting. None of these activities are related to her efforts on this project. Dr. Conti holds equity in Quintiles Health unrelated to her efforts on this project.

## Patient and public involvement

Patients and/or the public were not involved in the design, conduct, or reporting, or dissemination plans of this research. Patient consent for publication: Not applicable.

## Ethics approval

Not applicable.

## Data availability statement

All data used in writing this article are publicly available from sources referenced in the text. All extracted data is provided in supplemental materials.

## Notes

### Competing Interest Statement

The authors have declared no competing interest.

## REFERENCES

1. The selection and use of essential medicines 2023. World Health Organization 2023.

2. Farrow K. The Downstream Impacts of High Drug Costs for PrEP Have Hindered the Promise of HIV Prevention. Journal of Law, Medicine & Ethics. 2022;50(S1):47–50.

3. Roy V. Capitalizing a Cure: How Finance Controls the Price and Value of Medicines: University of California Press; 2023.

4. Mazzucato M, Roy V. Rethinking value in health innovation: from mystifications towards prescriptions. Journal of Economic Policy Reform. 2018;22(2):101–19.

5. Lazonick W, Tulum Ö, Hopkins M, Sakinç ME, Jacobson K. Financialization of the U.S. pharmaceutical industry. Institute for New Economic Thinking. 2019.

6. Tulum Ö, Lazonick W. Financialized Corporations in a National Innovation System: The U.S. Pharmaceutical Industry. International Journal of Political Economy. 2018;47(3-4):281–316.

7. Hatch O, Wyden R. THE PRICE OF SOVALDI AND ITS IMPACT ON THE U.S. HEALTH CARE SYSTEM. COMMITTEE ON FINANCE UNITED STATES SENATE 2015.

8. HIV PREVENTION DRUG: BILLIONS IN CORPORATE PROFITS AFTER MILLIONS IN TAXPAYER INVESTMENTS: Hearing before the House Oversight and Reform(5/16/2019, 2019).

9. Mitchell RK, Van Buren III HJ, Greenwood M, Freeman RE. Stakeholder Inclusion and Accounting for Stakeholders. 2015;52(7):851–77.

10. Lingane A, Olsen S. Guidelines for Social Return on Investment. California Review Managment. 2004;46(3):116–35.

11. Neumann PJ, Garrison LP, Willke RJ. The History and Future of the “ISPOR Value Flower”: Addressing Limitations of Conventional Cost-Effectiveness Analysis. Value in Health. 2022;25(4):558–65.

12. Lakdawalla DN, Doshi JA, Garrison Jr LP, Phelps CE, Basu A, Danzon PM. Defining Elements of Value in Health Care—A Health Economics Approach: An ISPOR Special Task Force Report [3]. Value in Health. 2018;21(2):131–9.

13. Stiglitz JE, Fitoussi J-P, Durand M. Measuring What Counts: The Global Movement for Well-Being: The New Press; 2019.

14. Christensen CM. The Innovator’s Dilemma: When New Technologies Cause Great Firms to Fail. Boston, Massachusetts: Harvard Business Review Press; 1997.

15. Phills Jr. JA, Deiglmeier K, Miller DT. Rediscovering Social Innovation. Stanford Social Innovation Review. 2008;6(4):34–43.

16. Sood N, Shih T, Van Nuys K, Goldman DP. Follow The Money: The Flow Of Funds In The Pharmaceutical Distribution System: Health Affairs Forefront; 2017 [Available from: 10.1377/forefront.20170613.060557.

17. 2020-2023 Value Assessment Framework. ICER; 2020.

18. Neumann PJ, Cohen JT, Weinstein MC. Updating Cost-Effectiveness — The Curious Resilience of the $50,000-per-QALY Threshold. N Engl J Med. 2014;371(9):796–7.

19. Weinstein MC, Torrance G, McGuire A. QALYs: The Basics. Value in Health. 2009;12, Supplement 1:S5–S9.

20. Cohen JT, Neumann PJ, Ollendorf DA. The much-maligned ‘quality-adjusted life year’ is a vital tool for health care policy2023. Available from: https://www.statnews.com/2023/03/22/the-much-maligned-quality-adjusted-life-year-is-a-vital-tool-for-health-care-policy/.

21. Kouakou CRC, Poder TG. Willingness to pay for a quality-adjusted life year: a systematic review with meta-regression. Eur J Health Econ. 2022;23(2):277–99.

22. Herman D, Afulani P, Coleman-Jensen A, Harrison GG. Food Insecurity and Cost-Related Medication Underuse Among Nonelderly Adults in a Nationally Representative Sample. Am J Public Health. 2015;105(10):e48–e59.

23. Berkowitz SA, Seligman HK, Choudhry NK. Treat or eat: food insecurity, cost-related medication underuse, and unmet needs. Am J Med. 2014;127(4):303-10.e3.

24. Zhou EW, Chaves da Silva PG, Quijada D, Ledley FD. Considering Returns on Federal Investment in the Negotiated “Maximum Fair Price” of Drugs Under the Inflation Reduction Act: an Analysis (Working Paper No. 219). Institute for New Economic Thinking. 2024.

25. Porter ME, Kramer MR. Creating Shared Value. Harvard Business Review. 2011;89, nos. 1-2:p62-77.

26. Statement on the Purpose of a Corporation. Business Roundtable; 2019.

27. Reyes-Urueña J, Campbell C, Diez E, Ortún V, Casabona J. Can we afford to offer pre-exposure prophylaxis to MSM in Catalonia? Cost-effectiveness analysis and budget impact assessment. AIDS Care. 2018;30(6):784–92.

28. Iino H, Hashiguchi M, Hori S. Estimating the range of incremental cost-effectiveness thresholds for healthcare based on willingness to pay and GDP per capita: A systematic review. PLoS One. 2022;17(4):e0266934.

29. 2020 U.S LIFE SCIENCES SALARY REPORT. BioSpace; 2020.

30. Sachan N, Tatambothla A, Nehru R, Dhanaraj C. Collaborative Commercialization at Gilead Sciences: Resolving the Innovation Vs. Access Tradeoff (Teaching Note). Harvard Business Impact; 2013.

31. Yang J, Qi J-L, Wang X-X, Li X-H, Jin R, Liu B-Y, et al. The burden of hepatitis C virus in the world, China, India, and the United States from 1990 to 2019. Front Public Health. 2023;2(11):1041201.

32. Wu J, Lai T, Han H, Liu J, Wang S, Lyu J. Global, regional and national disability-adjusted life years due to HIV from 1990 to 2019: findings from the Global Burden of Disease Study 2019. Trop Med Int Health. 2021;26(6):610–20.

33. Lakdawalla DN, Sun EC, Jena AB, Reyes CM, Goldman DP, Philipson TJ. An economic evaluation of the war on cancer. J Health Econ. 2010;29(3):333–46.

34. Philipson TJ, Jena AB. Who benefits from new medical technologies? Estimates of consumer and producer surpluses for HIV/AIDS drugs. Forum for Health Economics & Policy. 2006;9(2).

35. Moreno SG, Ray JA. The value of innovation under value-based pricing. J Mark Access Health Policy. 2016;7(4).

36. Camejo RR, McGrath C, Miraldo M, Rutten F. Distribution of health-related social surplus in pharmaceuticals: an estimation of consumer and producer surplus in the management of high blood lipids and COPD. Eur J Health Econ. 2014;15(4):439–45.

37. Grabner M, Johnson W, Abdulhalim AM, Kuznik A, Mullins CD. The Value of Atorvastatin Over the Product Life Cycle in the United States. Clinical Therapeutics. 2011;33(10):1433–43.

38. Lindgren P, Jönsson B. Cost-effectiveness of statins revisited: lessons learned about the value of innovation. Eur J Health Econ. 2012;13(4):445–50.

39. Jena AB, Philipson TJ. Cost-effectiveness analysis and innovation. J Health Econ. 2008;27(5):1224–36.

40. Garrison Jr. LP, Jiao B, Elsisi Z, Yehoshua A, Koruth R, Kreter B, et al. Estimating the Allocation of the Economic Value Generated by Utilization of All-Oral Direct-Acting Antivirals for Hepatitis C in the United States, 2015 to 2019. Value Health. 2024;27(8):1021–9.

41. Gilead Announces New License Agreement With the Medicines Patent Pool for Access to Bictegravir [press release]. Foster City, California: Gilead Sciences, Inc., October 04, 2017 2017.

42. Demeulemeester R, Savy N, Mounié M, Molinier L, Delpierre C, Dellamonica P, et al. Economic impact of generic antiretrovirals in France for HIV patients’ care: a simulation between 2019 and 2023. BMC Health Serv Res. 2022;22(1):567.

43. Sosnowy C, Predmore Z, Dean LT, Raifman J, Chu C, Galipeau D, et al. Paying for PrEP: A qualitative study of cost factors that impact pre-exposure prophylaxis uptake in the US. Int J STD AIDS. 2022;33(14):1199–205.

44. Cleary EG, Jackson MJ, Ledley FD. Government as the First Investor in Biopharmaceutical Innovation: Evidence From New Drug Approvals 2010–2019 (Working Paper No. 133). Institute for New Economic Thinking. 2020.

45. Cleary EG, Jackson MJ, Zhou EW, Ledley FD. Comparison of Research Spending on New Drug Approvals by the National Institutes of Health vs the Pharmaceutical Industry, 2010-2019. JAMA Health Forum. 2023;4(4):e230511.

46. Galkina Cleary E, Beierlein JM, Khanuja NS, McNamee LM, Ledley FD. Contribution of NIH funding to new drug approvals 2010–2016. Proc Natl Acad Sci USA. 2018;115(10):2329–34.

47. Rennane S, Baker L, Mulcahy A. Estimating the Cost of Industry Investment in Drug Research and Development: A Review of Methods and Results. Inquiry. 2021;58:00469580211059731.

48. DiMasi JA, Grabowski HG, Hansen RW. Innovation in the pharmaceutical industry: New estimates of R&D costs. J Health Econ. 2016;47:20–33.

49. Flier JS. Academia and industry: allocating credit for discovery and development of new therapies. J Clin Invest. 2019;129(6):2172–4.

50. Cleary EG, Jackson MJ, Folchman-Wagner Z, Ledley FD. Foundational research and NIH funding enabling Emergency Use Authorization of remdesivir for COVID-19. MedRxiv. 2020:2020.07. 01.20144576.

51. Barenie RE, Avorn J, Tessema FA, Kesselheim AS. Public funding for transformative drugs: the case of sofosbuvir. Drug Discov Today. 2021;26(1):273–81.

52. Johnson JA. AIDS Funding for Federal Government Programs: FY1981-FY2009. Congressional Research Service; 2008.

53. Edwards DJ, Coppens DG, Prasad TL, Rook LA, Iyer JK. Access to hepatitis C medicines. Bulletin of the World Health Organization. 2015;93(11):799–805.

54. Douglass CH, Pedrana A, Lazarus JV, ‘t Hoen EFM, Hammad R, Leite RB, et al. Pathways to ensure universal and affordable access to hepatitis C treatment. BMC Med. 2018;16(175).

55. Auty SG, Griffith KN, Shafer PR, Gee RE, Conti RM. Improving Access to High-Value, High-Cost Medicines: The Use of Subscription Models to Treat Hepatitis C using Direct Acting Antivirals in the United States. J Health Polit Policy Law. 2022;47(6):691–708.

56. Neumann PJ, Weinstein MC. Legislating against use of cost-effectiveness information. N Engl J Med. 2010;363(16):1495–7.

57. Rand LZ, Kesselheim AS. Controversy Over Using Quality-Adjusted Life-Years In Cost-Effectiveness Analyses: A Systematic Literature Review. Health Aff (Millwood). 2021;40(9):1402–10.

58. McDougall JA, Furnback WE, Wang BCM, Mahlich J. Understanding the global measurement of willingness to pay in health. J Mark Access Health Policy. 2020;8(1):1717030.

59. Martín-Fernández J, Polentinos-Castro E, Del Cura-González MI, Ariza-Cardiel G, Abraira V, Gil-Lacruz AI, et al. Willingness to pay for a quality-adjusted life year: an evaluation of attitudes towards risk and preferences. BMC Health Serv Res. 2014;14(287).

60. SEC Staff Accounting Bulletin: No. 99 – Materiality. U.S. Securities and Exchange Commision; 1999.

61. Huang Y-lA, Zhu W, Smith DK, Harris N, Hoover KW. HIV Preexposure Prophylaxis, by Race and Ethnicity — United States, 2014–2016. MMWR Morb Mortal Wkly Rep. 2018;67(41):1147–50.

62. National Center for HIV, Viral Hepatitis, STD, and Tuberculosis Prevention: About AtlasPlus: CDC; 2024 [Available from: https://www.cdc.gov/nchhstp/about/atlasplus.html.

