## Supplemental Materials for "Social and private value created through commercialization of Gilead Sciences’ innovative medicines for Hepatitis C and HIV: a cross-sectional analysis"

**Supplemental methods 1**. Data Sources, Differences Between the Use of QALYs in Our Study versus in Cost-Effectiveness Analysis, Exclusion Criteria, and Definitions of Accounting and Finance Terms

**Supplemental table 1**. Median Gains in Quality-Adjusted Life Years (QALYs) for Each of Gilead's Leading Products

**Supplemental table 2**. Total Health Value Created by Gilead's Leading Medicines (2012-2020) Using US WTP/QALY ($104K)

**Supplemental table 3**. Gilead Medicines Commercialized for HCV and HIV 2012-2020

**Supplemental table 4**. Gilead Products and Product Revenue 2012-2020

**Supplemental table 5**. Residual Health Value Created by Gilead's Leading Medicines (2012-2020) Using US WTP/QALY ($104K)

**Supplemental table 6**. Financial Statements of Gilead Sciences 2012-2020

**Supplemental figure 1**. Gilead's Select HCV and HIV Product Revenue 2012-2020

**Supplemental figure 2**. Summary of Gilead Financial Data and Metrics 2012-2020

**Supplemental figure 3**. Social and Private Value Creation by Gilead 2012-2020

**eReferences.**

**Supplemental methods 1. Data Sources,** **Differences Between the Use of QALYs in Our Study versus in Cost-Effectiveness Analysis, Exclusion Criteria, and Definitions of Accounting and Finance Terms.**

***Data Sources***

Gilead’s product-specific revenue 2012-2020 were from SEC filings. HCV and HIV products accounted for >3% of Gilead revenue over this period and were included in this analysis (supplemental table 4). Gilead reports revenue net of rebates.

Publications reporting QALYs gained were identified in the Cost-Effectiveness Analysis (CEA) Registry ([www.cearegistry.org](http://www.cearegistry.org), accessed October 2021)^1^ or PubMed (<https://pubmed.ncbi.nlm.nih.gov>, accessed October 2021). Units sold and wholesale price paid including out-of-pocket and third-party costs for branded, unbranded, or authorized generic products, net prompt pay discounts and chargebacks, but not net rebates paid after the fact, were from IQVIA MIDAS® (accessed August 2021). Data was obtained for the US and 77 countries in global (ex-US) markets. The fraction of individuals using Truvada or Descovy for PrEP versus therapy through 2016 was from Morbidity Mortality Weekly Report^2^ and from 2017-2020 was from the National Center for HIV, Viral Hepatitis, STD, and TB Prevention, AtlasPlus (https://www.cdc.gov/nchhstp/about/atlasplus.html). Average full-time salaries were from BioSpace 2020 US Life Sciences Salary report.^3^ Government fines were from Violation Tracker (https://violationtracker.goodjobsfirst.org).^4^ Audited financial data was from Compustat (WRDS, <https://wrds-www.wharton.upenn.edu>). Definitions of Compustat terms are found below in the “Definitions of Accounting and Finance Terms” section of this supplemental methods.

Analyses were done in Microsoft Excel version 2311.

***Differences Between the Use of QALYs in Our Study versus in Cost-Effectiveness Analysis***

While this work uses QALY units as a measure of health value, there are essential differences in our use of this metric and its application in cost-effectiveness studies.^5-7^ First, the present study measures health value as total QALYs gained relative to untreated controls rather than incremental QALYs gained over alternative therapies.^8^ As such, our analysis focuses explicitly on the total health value accruing to an individual using the product. This analysis does not include health care costs. Second, we use willingness-to-pay (WTP) to model the economic value of a QALY, rather than threshold values reflecting broader opportunity costs to society. Finally, this analysis considered residual health value (net price paid) – not total health value - to be the most appropriate measure of health benefit accruing to those treated with the product. This reflects evidence that high drug prices are associated with poor compliance of prescribed treatment regiments as well as greater economic, food, and housing insecurity, which may collectively compromise the ability of individuals to realize the full benefit of these medicines.^9-11^

***Exclusion Criteria***

The exclusion criteria for publications reporting QALYs were as follows: review papers; studies focusing on surgical interventions (e.g., transplants) or non-pharmaceutical approaches (e.g., treatment timing, screening methods, strategic plans, or care models); studies not specifying the direct-acting antiviral (DAA) therapy used; studies involving drug combinations of Gilead and non-Gilead products (e.g., Sofosbuvir and Simeprevir); studies not reporting total QALYs; non-English-language publications; and studies published before the approval date of each drug OR published after 2021.

***Definitions of Accounting and Finance Terms***

- COGS = Cost of Goods Sold
- CSHO = Common Shares Outstanding
- DP = Depreciation and Amortization
- DVT = Dividends - Total
- EBITDA = Earnings Before Interest, Taxes, Depreciation, and Amortization
- EMP = Employees
- GP = Gross Profit
- NI = Net Income (Loss)
- PRCC_F = Closing Price at Fiscal Year-End
- PRSTKC = Purchase of Common and Preferred Stock
- RDIP = In-Process R&D
- REVT = Revenue
- TXT = Income Taxes - Total
- XRD = Research & Development (R&D) Expense
- XSGA = Selling, General & Administrative (SG&A) Expense

*Cost of Goods Sold – Compustat variable = COGS, equivalent to REVT - GP*

COGS represents all costs related to producing or purchasing the products sold during the year. For pharmaceutical companies, it includes manufacturing costs for the product (i.e., drug) sold during the year, covering, for example, raw materials, formulation, packaging, labor costs associated with production and distribution, and the cost of Quality Control, Quality Assessment, and FDA compliance as well as the allocated overhead costs associated with operating production facilities, personnel, and related services.

In this study, COGS was summed to XSGA and DP (and non-Gilead drug costs) to estimate the network value.

*Depreciation and Amortization – Compustat variable = DP*

DP reflects the value of business assets over time. Depreciation allocates the cost of physical assets (e.g., machinery and buildings) over time to match expenses with generated revenue. Amortization spreads the cost of intangible assets (patents, copyrights, trademarks) over their estimated useful lives.

In this study, we included DP along with COGS and SG&A (and non-Gilead drug costs) to estimate the network value.

*Distributions to Shareholders - Total - Calculated – Compustat variables = DVT + PRSTKC*

This study used the total distribution to shareholders to estimate the shareholder value.

*Earnings Before Interest, Taxes, Depreciation, and Amortization – Compustat variable = EBITDA*

EBITDA is used to evaluate a company's operating performance, excluding the indebtedness, state-mandated payments, and expenses necessary to sustain its assets.

In this study, EBITDA was included in supplemental table 6 to exhibit Gilead’s overall financial statement.

*Employees – Compustat variable = EMP*

EMP represents the company’s reported number of workers to shareholders. Some firms report it as an average, while others provide the year-end count, with preference given to the latter when both are available. EMP includes all employees of consolidated subsidiaries (both domestic and foreign). EMP excludes consultants, contract workers, directors, and employees of unconsolidated subsidiaries.

In this study, EMP was used to estimate job creation, when multiplied by average salaries in the biotechnology sector.

*Gross Profit – Compustat variable = GP, equivalent to REVT - COGS*

GP reflects the difference between REVT and COGS before any deduction of the other costs of running a company such as R&D or XSGA.

In this study, GP was included in supplemental table 6 to exhibit Gilead’s overall financial statement.

*Income Taxes - Total – Compustat variable = TXT*

TXT is a tax on the company profits. In Compustat, it represents all income taxes imposed by federal, state, and foreign governments.

In this study, we used TXT as one of the components of social value, and it was added to fines and other payments to the public sector (collected from Violation Tracker^1^).

*Market Capitalization - Calculated – Compustat variables = CSHO x PRCC_F*

Market Capitalization refers to the total market value of a company’s outstanding shares, calculated as the number of common shares outstanding multiplied by the company’s stock price. Market Capitalization is a proxy for company size.

In this study, Market Capitalization was utilized to calculate shareholder value (see below).

Change in Market Capitalization = Market Capitalization in year y minus Market Capitalization in year y-1

*Net Income – Compustat variable = Net Income (Loss) NI*

NI represents the fiscal period income or loss reported by a company after subtracting expenses and losses from all revenues and gains.

In this study, NI was included in supplemental table 6 to exhibit Gilead’s overall financial statement.

*Purchase of Common and Preferred Stock – Compustat variable = PRSTKC*

PRSTKC represents any use of funds decreasing common and/or preferred stock.

In this study, PRSTKC was used to estimate the shareholder value.

*Research & Development (R&D) Expense - Calculated – Compustat variables = XRD + RDIP*

XRD includes basic, applied, and translational research, process development, and pilot production, and research required for regulatory approval,^12^ but specifically excludes the costs associated with commercial production, quality control, ongoing product improvements, marketing research, and costs associated with patent applications or litigation.^13^

RDIP represents the R&D “purchased” through the transaction. Compustat includes R&D expense and in-process R&D in the variable XRD. RDIP is coded as a negative number in Compustat. Therefore, to get R&D expense without in-process R&D, RDIP is added to XRD.

In this study, XRD + RDIP was used to estimate the scientific value.

*R&D Intensity - Calculated – Compustat variables = XRD + RDIP / REVT*

All margins were calculated with REVT as the denominator.

*Revenue – Compustat variable = REVT*

REVT represents the total amount of actual billings to customers for regular sales, reduced by cash discounts, trade discounts, and returned sales and allowances for which credit is given to customers.

In this study, revenue data was utilized to identify Gilead's most profitable antiviral products for analysis (supplemental table 4), excluding those contributing less than 3% of total revenue (2012-2020), as well as the peak sales per drug (supplemental figure 1).

*Selling, General & Administrative (SG&A) Expense - Calculated – Compustat variables = XSGA - XRD*

XSGA includes expenses related to revenue generation (e.g., marketing, advertising, delivery) or in operational activities not directly tied to production. XSGA represents the cost of corporate operations such as marketing and sales, human resources, facilities (not part of manufacturing), and management. The Compustat variable XSGA includes XRD in most cases. To avoid double counting, we subtract XRD from XSGA.

In this study, XSGA was summed to COGS and DP (and non-Gilead drug costs) to estimate the network value.

*Shareholder Value*

Change in Market Capitalization + DVT + Stock Buybacks

**Supplemental Table 1. Median Gains in Quality-Adjusted Life Years (QALYs) for Each of Gilead's Leading Products.**

| **Product** | **PMID^a^** | **Indication** | **QALY Unit^b^** | **Avg. QALYs  (Untreated)^c^** | **Avg. QALYs  (Treated)^d^** | **QALYs Gained^e^** |
| --- | --- | --- | --- | --- | --- | --- |
| Sovaldi | 26990023 | HCV | lifetime - per person | 17.16 | 20.07 | 2.91 |
|  | 29756049 | HCV | lifetime - per person | 8.93 | 10.97 | 2.04 |
|  | 28401125 | HCV | lifetime - per person | 8.20 | 10.80 | 2.60 |
|  | 27342742 | HCV | lifetime - per person | 9.15 | 12.05 | 2.90 |
|  | 25619871 | HCV | lifetime - per person | 12.19 | 14.80 | 2.61 |
|  | 25974722 | HCV | lifetime - per person | 11.4 | 12.95 | 1.55 |
|  | 25329202 | HCV | lifetime - per person | 13.17 | 15.25 | 2.08 |
|  | **Median Lifetime QALYs Gained per Person** | | | | | ***2.60*** |
| Harvoni | 26990023 | HCV | lifetime - per person | 17.16 | 20.42 | 3.26 |
|  | 29756049 | HCV | lifetime - per person | 8.93 | 11.24 | 2.31 |
|  | 28401125 | HCV | lifetime - per person | 8.20 | 10.56 | 2.36 |
|  | 28520728 | HCV | lifetime - per person | 12.71 | 17.30 | 4.59 |
|  | 31808054 | HCV | lifetime - per person | 10.19 | 14.20 | 4.01 |
|  | 29864213 | HCV | lifetime - per person | 14.42 | 17.00 | 2.58 |
|  | 27342742 | HCV | lifetime - per person | 9.15 | 13.36 | 4.21 |
|  | 27063573 | HCV | lifetime - per person | 13.54 | 16.39 | 2.85 |
|  | 25619871 | HCV | lifetime - per person | 12.19 | 15.86 | 3.67 |
|  | **Median Lifetime QALYs Gained per Person** | | | | | ***3.26*** |
| Epclusa | 29864213 | HCV | lifetime - per person | 14.42 | 18.36 | 3.94 |
|  | **Median Lifetime QALYs Gained per Person** | | | | | ***3.94*** |
| Atripla^f^ | 22163167 | HIV (non-PrEP) | Annual - per person | 0.525 | 0.76 | 0.23 |
|  | 23028230 | HIV (non-PrEP) | Annual - per person | 0.790 | 0.98 | 0.19 |
|  | 23430273 | HIV (non-PrEP) | Annual - per person | 0.525 | 0.76 | 0.23 |
|  | **Median Annual QALYs Gained per Person** | | | | | ***0.23*** |
| Truvada^g^ | 29262694 | HIV (PrEP) | lifetime - per person | 20.1 | 24.30 | 4.19 |
|  | 27110953 | HIV (PrEP) | lifetime - population | 6.434^h^ | 6.4341^h^ | 5.80^i^ |
|  | 25798150 | HIV (PrEP) | lifetime - per person | 20.21 | 25.73 | 5.53 |
|  | **Median Lifetime QALYs Gained per Person** | | | | | ***5.53*** |
|  | **Median Annual QALYs Gained per Person** | | | | | ***0.35*^j^** |

Publications reporting QALYs were identified in the CEA Registry^1^ searching for “QALY,” [title and abstract fields], “cost-effectiveness,” [title and abstract fields], filtering by intervention [pharmaceutical intervention], and indication [Certain infections and parasitic diseases (A00-B99)]. PubMed (accessed October 2021) search: brand or generic name and QALY. Papers preceding the drug approval were excluded. ^a^PubMed ID associated with the publication or study for the specific product. ^b^Unit of measurement for QALYs. ^c^Average QALYs per person for non-treatment. ^d^Average QALYs per person after treatment. ^e^Difference between Avg. QALYs (Treated) and Avg. QALYs (Untreated). ^f^Studies with annual QALYs per person were prioritized to avoid QALY conversion. ^g^QALYs gained refers to QALYs gained per infection averted. ^h^In billions. ^i^Individual QALYs converted from HIV infection averted. ^j^Annual QALYs converted from lifetime QALYs, considering 20-year time horizon and 3% discount rate.

**Supplemental Table 2. Total Health Value Created by Gilead's Leading Medicines (2012-2020) Using US WTP/QALY ($104K).**

|  |  | **US Population** | | | **Global Population (ex-US)** | | |
| --- | --- | --- | --- | --- | --- | --- | --- |
| **Product** | **QALYs**  **Gained** | **# Individuals Benefited** | **Health Benefit (QALYs)** | **Health Value (billions)** | **# Individuals Benefited** | **Health Benefit (QALYs)** | **Health Value (billions)** |
| ***HCV*** | | | | | | | |
| Sovaldi | 2.60 | 156,655 | 407,304 | $42.4 | 1,640,750 | 4,265,951 | $443.7 |
| Harvoni | 3.26 | 388,105 | 1,265,222 | $131.6 | 468,633 | 1,527,744 | $158.9 |
| Epclusa | 3.94 | 176,332 | 694,750 | $72.3 | 574,693 | 2,264,291 | $235.5 |
| **Total HCV** |  | **721,093** | **2,367,276** | **$246.2** | **2,684,077** | **8,057,987** | **$838.0** |
| ***HIV*** | | | | | | | |
| Truvada (non-PrEP) | 0.23 | 1,394,585 | 320,754 | $33.4 | 2,296,898 | 528,287 | $54.9 |
| Truvada (PrEP) | 0.35 | 13,044 | 4,565 | $0.48 | 25,780 | 9,023 | $0.94 |
| Atripla | 0.23 | 1,104,181 | 253,962 | $26.4 | 12,371,245 | 2,845,386 | $295.9 |
| Stribild | 0.23 | 322,717 | 74,225 | $7.7 | 173,539 | 39,914 | $4.2 |
| Genvoya | 0.23 | 693,525 | 159,511 | $16.6 | 445,072 | 102,367 | $10.6 |
| Biktarvy | 0.23 | 475,442 | 109,352 | $11.4 | 307,998 | 70,839 | $7.4 |
| Descovy (non-PrEP) | 0.23 | 333,077 | 76,608 | $8.0 | 283,795 | 65,273 | $6.8 |
| Descovy (PrEP) | 0.35 | 2,688 | 941 | $0.98 | 2,058 | 720 | $0.75 |
| **Total HIV** |  | **4,339,258** | **999,917** | **$104.0** | **15,906,384** | **3,661,809** | **$380.8** |
| **TOTAL** |  | **5,781,443** | **5,734,469** | **$350.2** | **21,274,538** | **19,777,783** | **$1,218.9** |

All monetary values in USD (billions) inflation was adjusted to 2016. # individuals treated in US and global (ex-us) population was from MIDAS. Health Value was calculated with an adjusted US WTP/QALY of $104,000.^14^ The fraction of individuals treated with Truvada or Descovy for PrEP was from estimates in Morbidity Mortality Weekly Report^15^ through 2016 or the National Center for HIV, Viral Hepatitis, STD, and TB Prevention from 2017-2020. https://www.cdc.gov/nchhstp/default.htm [Values used: 2012=0%, 2013=0%, 2014=6%, 2015=14%, 2016=28%, 2017=63%, 2018=85%, 2019=69%, 2020=69%. The number of individuals who benefited from PrEP is divided by the Number Needed to Treat (NNT) of 58.1.^16^

**Supplemental Table 3. Gilead Medicines Commercialized for HCV and HIV 2012-2020.**

| **Brand Name** | **Generic Name** | **Acronym** |
| --- | --- | --- |
| ***HCV*** | | |
| Sovaldi | Sofosbuvir | SOF |
| Harvoni | Sofosbuvir/Ledipasvir | SOF/LDV |
| Epclusa | Sofosbuvir/Velpatasvir | SOF/VEL |
| ***HIV*** | | |
| Truvada | Emtricitabine/Tenofovir Disoproxil Fumarate | FTC/TDF |
| Atripla | Efavirenz/Emtricitabine/Tenofovir Disoproxil Fumarate | EFV/FTC/TDF |
| Stribild | Elvitegravir/Cobicistat/Emtricitabine/Tenofovir Disoproxil Fumarate | EVG/COBI/FTC/TDF |
| Genvoya | Elvitegravir/Cobicistat/Emtricitabine/Tenofovir alafenamide | EVG/COBI/FTC/TAF |
| Biktarvy | Bictegravir/Emtricitabine/Tenofovir Alafenamide | BIC/FTC/TAF |
| Descovy | Emtricitabine/Tenofovir Alafenamide | FTC/TAF |

**Supplemental table 4. Gilead Products and Product Revenue 2012-2020.**

| **Product** | **FDA Approval** | **Cumulative Revenue** | **% of Total Revenue** |
| --- | --- | --- | --- |
| ***HCV*** | | | |
| Sovaldi | 1/13 | $20.9 | 10.4% |
| Harvoni | 10/14 | $31.6 | 15.7% |
| Epclusa | 6/16 | $10.4 | 5.2% |
| ***HIV*** | | | |
| Truvada | 08/04 (non-PrEP) 07/12 (PrEP) | $26.8 | 13.3% |
| Atripla | 12/6 | $25.6 | 12.7% |
| Stribild | 8/12 | $7.8 | 3.9% |
| Genvoya | 11/15 | $15.1 | 7.5% |
| Biktarvy | 2/18 | $11.8 | 5.9% |
| Descovy | 04/16 (non-PrEP) 10/19 (PrEP) | $6.1 | 3.1% |
| **TOTAL** |  | **$156.0** | **77.6%** |

All values for years in USD (billions) and inflation adjusted to 2016. Gilead reports revenue net of rebates.

**Supplemental table 5. Residual Health Value Created by Gilead's Leading Medicines (2012-2020) Using US WTP/QALY ($104K).**

| **Generic name** | **Total US price paid** | **US Residual Health Value** | **Total Global  (ex-US) price paid** | **Global Residual Health Value** |
| --- | --- | --- | --- | --- |
| ***HCV*** | | | | |
| Sovaldi | $13.1 | $29.3 | $13.0 | $430.7 |
| Harvoni | $33.4 | $98.1 | $15.9 | $142.9 |
| Epclusa | $9.0 | $63.3 | $10.9 | $224.6 |
| **Total HCV** | **$55.5** | **$190.7** | **$39.8** | **$798.2** |
| ***HIV*** | | | | |
| Truvada | $22.2 | $11.2 | $11.3 | $44.6 |
| Atripla | $17.3 | $9.2 | $8.8 | $287.1 |
| Stribild | $6.7 | $1.1 | $1.5 | $2.7 |
| Genvoya | $14.8 | $1.8 | $3.5 | $7.2 |
| Biktarvy | $10.5 | $0.83 | $1.4 | $6.0 |
| Descovy | $5.2 | $2.8 | $1.8 | $5.1 |
| **Total HIV** | **$76.6** | **$26.8** | **$28.2** | **$352.6** |
| **TOTAL** | **$132.2** | **$217.4** | **$68.1** | **$1,150.8** |

Data source. Market Information Data Analytics System (MIDAS), Centers for Medicare & Medicaid Services (CMS), and AtlasPlus Centers for Disease Control and Prevention (CDC). All values in USD (billions), inflation was adjusted to 2016. Health Value range based on US WTP/QALY ($104,000).^14^ Price paid: wholesale price paid net of prompt pay discounts and chargebacks. US Residual Health Value: the net health value calculated as the difference between the total health value generated by the product and its spending within the US. The same calculation applies to Global Residual Health Value (excluding the United States). A positive value indicates a net health gain, while a negative value suggests a net health loss.

**Supplemental figure 1. Gilead's Select HCV and HIV Product Revenue 2012-2020.** A. HCV Product Sales by Year. B. HIV Product Sales by Year.

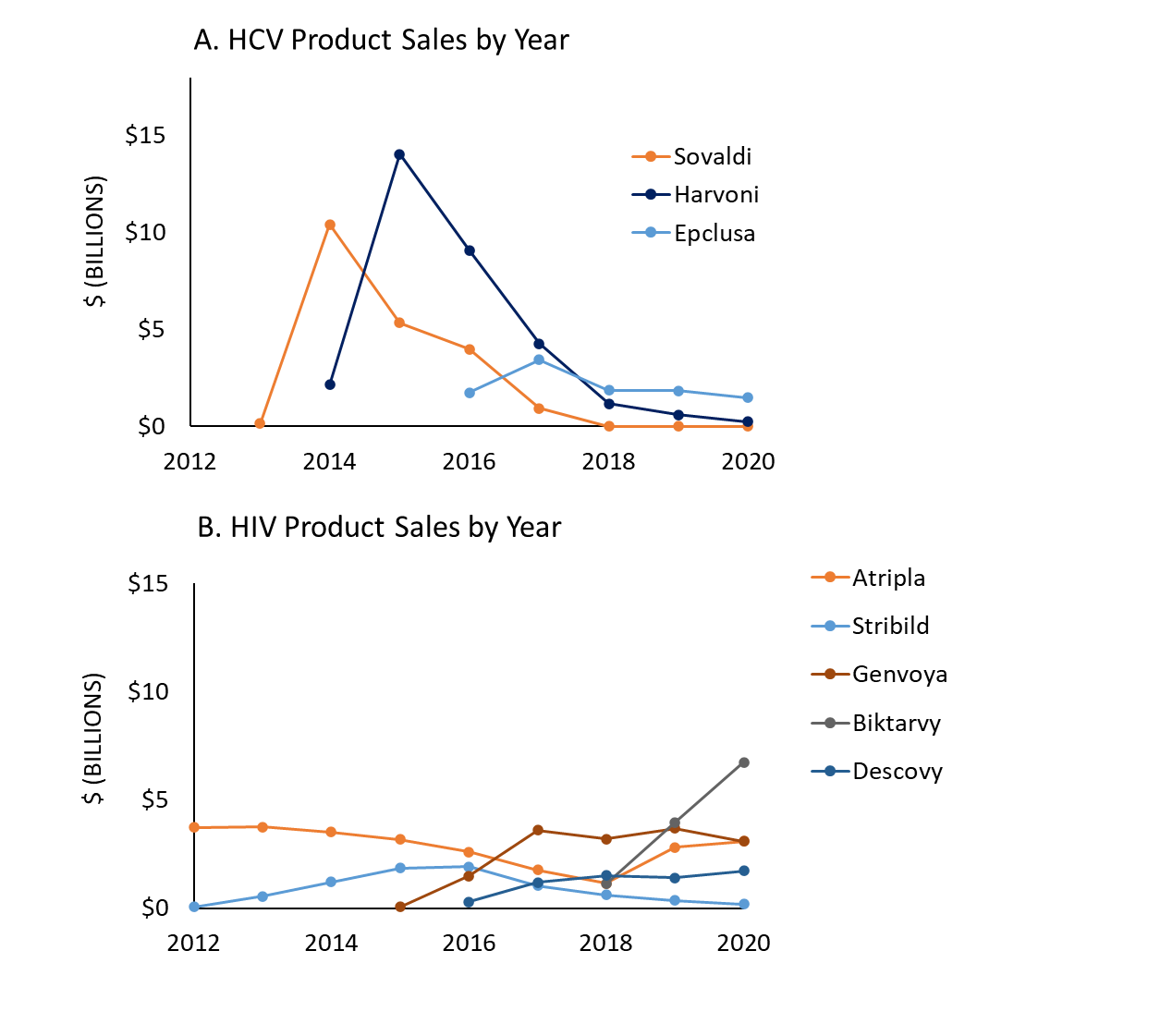

Data source: Gilead financial statement, 10K. All values in USD, billions; inflation was adjusted to 2016. A. Gilead revenue from HCV products: Sovaldi, Harvoni, Epclusa. After 2018, Sovaldi was not specified in Gilead’s revenue but reported as grouped with other products. B. Gilead revenue from HIV products: Truvada, Atripla, Stribild, Genvoya, Biktarvy, Descovy.

**Supplemental table 6. Financial Statements of Gilead Sciences 2012-2020.**

| *in billions* | **2012** | **2013** | **2014** | **2015** | **2016** | **2017** | **2018** | **2019** | **2020** | ***Total*** |
| --- | --- | --- | --- | --- | --- | --- | --- | --- | --- | --- |
| ***Basic Financial Numbers*** | | | | | | | | | | |
| Revenue - Total | $10.1 | $11.5 | $25.2 | $33.1 | $30.4 | $25.6 | $21.1 | $21.1 | $22.9 | **$201.0** |
| Gross Profit (Loss) | $7.7 | $8.8 | $22.3 | $30.0 | $27.2 | $22.4 | $17.9 | $18.0 | $20.1 | **$174.4** |
| EBITDA | $4.7 | $5.0 | $16.6 | $23.5 | $19.5 | $15.3 | $10.0 | $6.1 | $11.2 | **$111.8** |
| Net Income (Loss) | $2.7 | $3.2 | $12.3 | $18.3 | $13.5 | $4.5 | $5.2 | $5.1 | $0.1 | **$64.9** |
| R&D Expense | $1.8 | $2.2 | $2.9 | $3.1 | $4.3 | $3.4 | $4.0 | $7.8 | $4.7 | **$34.2** |
| SG&A Expense | $1.2 | $1.7 | $2.9 | $3.4 | $3.4 | $3.7 | $3.8 | $4.1 | $4.2 | **$28.5** |
| Cost of Goods Sold | $2.4 | $2.7 | $2.9 | $3.1 | $3.2 | $3.2 | $3.3 | $3.1 | $2.8 | **$26.6** |
| Income Taxes - Total | $1.1 | $1.2 | $2.8 | $3.6 | $3.6 | $8.7 | $2.2 | -$0.2 | $1.5 | **$24.5** |
| Job creation | $0.6 | $0.8 | $0.9 | $1.0 | $1.1 | $1.2 | $1.3 | $1.3 | $1.5 | **$9.7^a^** |
| ***Margins^b^*** | | | | | | | | | | |
| R&D intensity | 18.1% | 18.9% | 11.5% | 9.2% | 14.0% | 13.5% | 19.0% | 37.0% | 20.4% |  |
| Gross profit margin | 76.0% | 76.7% | 88.6% | 90.8% | 89.3% | 87.6% | 84.5% | 85.4% | 87.7% |  |
| Net income margin | 26.7% | 27.4% | 48.6% | 55.5% | 44.4% | 17.7% | 24.7% | 24.0% | 0.5% |  |
| Income Taxes margin | 10.7% | 10.3% | 11.2% | 10.9% | 11.9% | 34.0% | 10.6% | -0.9% | 6.4% |  |
| ***Stock Sales and Distributions*** | | | | | | | | | | |
| Market Cap | $58.3 | $118.7 | $143.2 | $145.7 | $93.8 | $91.8 | $76.6 | $77.2 | $67.7 | **$873.2** |
| Annual Change Market Cap | $25.4 | $60.4 | $24.5 | $2.5 | -$51.9 | -$2.1 | -$15.1 | $0.6 | -$9.5 | **$34.9** |
| Total distributions to shareholders | $0.7 | $0.6 | $5.4 | $12.0 | $13.5 | $3.6 | $5.6 | $4.7 | $4.7 | **$50.8** |
| Dividends - Total | $0.0 | $0.0 | $0.0 | $1.9 | $2.5 | $2.7 | $2.9 | $3.0 | $3.2 | **$16.2** |
| Purchase of Common and Preferred Stock | $0.7 | $0.6 | $5.4 | $10.1 | $11.0 | $0.9 | $2.8 | $1.6 | $1.5 | **$34.7** |
| Annual change shareholder value | $26.1 | $61.0 | $29.9 | $14.5 | -$38.4 | $1.6 | -$9.5 | $5.3 | -$4.8 | **$85.7** |

Data source: Compustat. All values in USD, billions; inflation was adjusted to 2016. EBITDA = Earnings Before Interest, Taxes, Depreciation, and Amortization. ^a^Based on the sum of employees 2012-2020 (81,500). ^b^Margins and R&D intensity were calculated on revenue. Revenue is reduced by discounts and chargebacks.

**Supplemental figure 2. Summary of Gilead Financial Data and Metrics 2012-2020.** A. Revenue and Net Income. B. Market Capitalization and Shareholder Value. C. Expense Metrics.

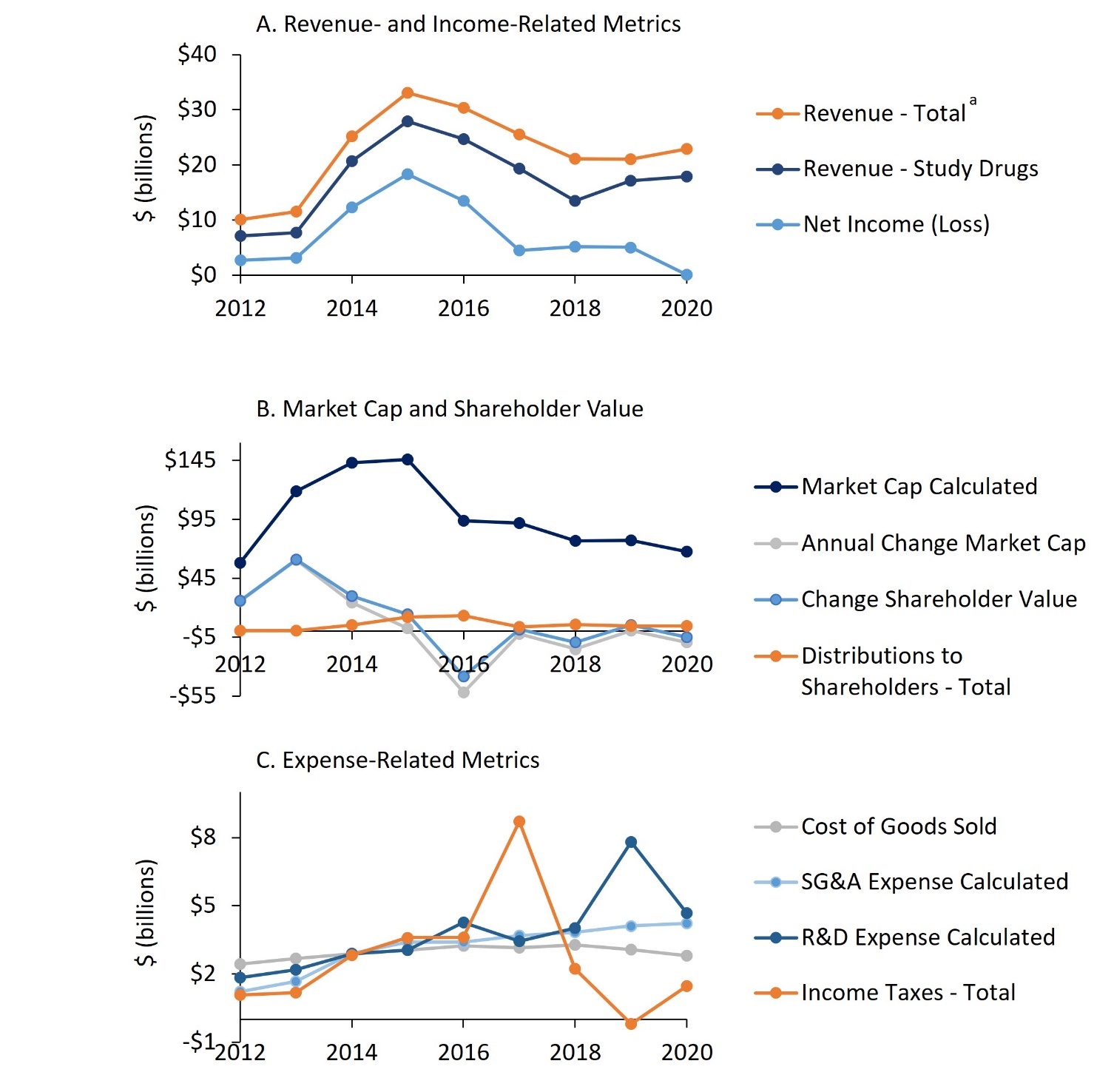

Data source: Compustat. All values in USD, billions; inflation was adjusted to 2016. A. Revenue and net income. Revenue is reduced by discounts and chargebacks. B. Total distributions to shareholders, market capitalization, change shareholder value, annual change market capitalization. Market capitalization= Common shares outstanding multiplied by price of stock. Change in market capitalization= market capitalization in year y minus market capitalization in year y-1. Shareholder value = change in market capitalization + dividends + stock buybacks. C. R&D expense calculated, cost of goods sold (COGS), income taxes, and SG&A. R&D expense calculated = R&D expense - In-Process R&D. SG&A calculated = Gross profit (loss) - EBITDA - R&D expense calculated.

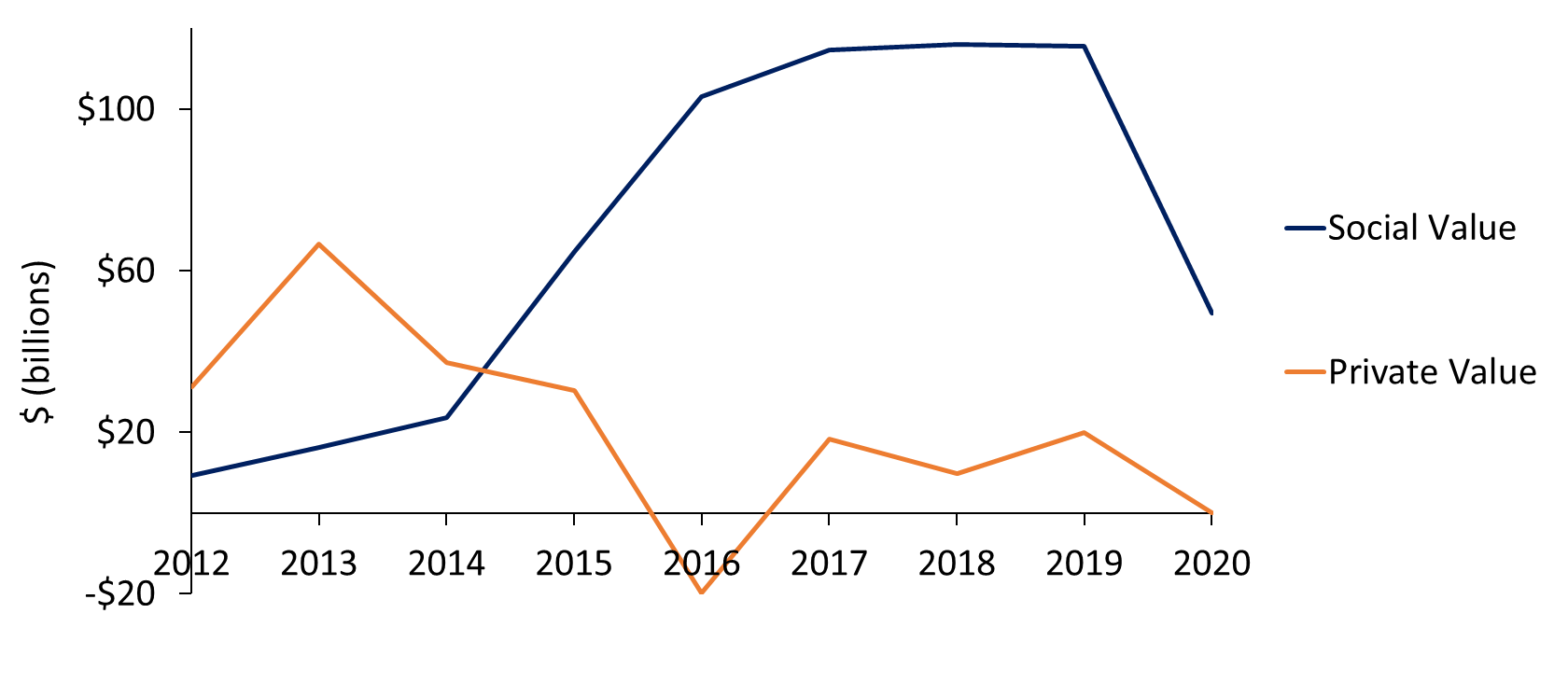
**Supplemental figure 3. Social and Private Value Creation by Gilead 2012-2020.**

All values in USD, billions; inflation was adjusted to 2016. Excludes fines ($97 million) and research collaboration ($453.5 million) due to their sporadic nature.

**eReferences.**

1. Center for the Evaluation of Value and Risk in Health. The Cost-Effectiveness Analysis Registry [Internet]. (Boston), Institute for Clinical Research and Health Policy Studies, Tufts Medical Center. Available from: www.cearegistry.org. Accessed in October 2021.
2. Huang YA, Zhu W, Smith DK, Harris N, Hoover KW. HIV preexposure prophylaxis by race and ethnicity 2014-2016. Vol. 67. 2018:1147-1150. US Department of Health and Human Services/Centers for Disease Control and Prevention.
3. REPORT USLSS. 2020 U.S. LIFE SCIENCES SALARY REPORT.
4. Violation Tracker. Corporate Research Project of Good Jobs First. <https://violationtracker.goodjobsfirst.org>
5. ICER. 2020-2023 Value Assessment Framework. 2020. Value Assessment Framework.
6. Neumann PJ, Cohen JT, Weinstein MC. Updating cost-effectiveness--the curious resilience of the $50,000-per-QALY threshold. N Engl J Med. Aug 28 2014;371(9):796-7. doi:10.1056/NEJMp1405158
7. Weinstein MC, Torrance G, McGuire A. QALYs: The Basics. Value in Health. 2009;12:S5-S9. doi:10.1111/j.1524-4733.2009.00515.x
8. Neumann PJ, Ganiats TG, Russell LB, Sanders GD, Siegel JE. Cost-Effectiveness in Health and Medicine. Oxford University Press; 2016.
9. Herman D, Afulani P, Coleman-Jensen A, Harrison GG. Food insecurity and cost-related medication underuse among nonelderly adults in a nationally representative sample. American journal of public health. 2015;105(10):e48-e59.
10. Berkowitz SA, Seligman HK, Choudhry NK. Treat or eat: food insecurity, cost-related medication underuse, and unmet needs. The American journal of medicine. 2014;127(4):303-310. e3.
11. Zhou EW, Chaves da Silva PG, Quijada D, Ledley FD. Considering Returns on Federal Investment in the Negotiated “Maximum Fair Price” of Drugs Under the Inflation Reduction Act: an Analysis (Working Paper No. 2019). Institute for New Economic Thinking. 2024;doi:https://doi.org/10.36687/inetwp219
12. FASB Accounting Standards Codification. 730-10-55-1. Research and Development – Implementation Guidance - Examples of Activities Typically Included in Research and Development. https://asc.fasb.org/1943274/2147483094/730-10-55-1
13. FASB Accounting Standards Codification. 730-10-55-2. Research and Development – Implementation Guidance - Examples of Activities Typically Excluded in Research and Development. <https://asc.fasb.org/1943274/2147483094/730-10-55-2>
14. Iino H, Hashiguchi M, Hori S. Estimating the range of incremental cost-effectiveness thresholds for healthcare based on willingness to pay and GDP per capita: a systematic review. PloS one. 2022;17(4):e0266934.
15. Huang YA, Zhu W, Smith DK, Harris N, Hoover KW. HIV preexposure prophylaxis by race and ethnicity 2014-2016. Vol. 67. 2018:1147–1150. US Department of Health and Human Services/Centers for Disease Control and Prevention.
16. Reyes-Urueña J, Campbell C, Diez E, Ortún V, Casabona J. Can we afford to offer pre-exposure prophylaxis to MSM in Catalonia? Cost-effectiveness analysis and budget impact assessment. AIDS Care. 2018/06/03 2018;30(6):784–792. doi:10.1080/09540121.2017.1417528
